# The proportion of Alzheimer’s disease attributable to apolipoprotein E

**DOI:** 10.1101/2023.11.16.23298475

**Authors:** Dylan M. Williams, Sami Heikkinen, Mikko Hiltunen, FinnGen, Neil M. Davies, Emma L. Anderson

## Abstract

Variation in the *APOE* gene strongly affects Alzheimer’s disease (AD) risk. However, the proportion of AD burden attributable to this variation requires clarification. We estimated the extents to which clinically diagnosed AD, AD neuropathology and all-cause dementia are attributable to the common *APOE* alleles in four large studies. First, we used data on 171,105 and 289,150 participants aged ≥60 years from UK Biobank (UKB) and FinnGen, respectively. AD and all-cause dementia were ascertained from linked electronic health records in these cohorts. Second, we examined amyloid-β positivity from amyloid positron emission tomography scans in 4,440 participants of the A4 Study. Third, we analysed data from the Alzheimer’s Disease Genetics Consortium (ADGC), where neuropathologically-confirmed AD cases were compared to pathology-negative, cognitively intact controls (N=5,007). In each analysis, we estimated outcome risk among carriers of *APOE* risk alleles ε3 and ε4, relative to individuals with an ε2/ε2 genotype, and calculated attributable fractions to show the proportions of the outcomes due to ε3 and ε4. For AD, fractions ranged from 71.5% (95% CI: 54.9%, 81.7%) in FinnGen to 92.7% in the ADGC (82.4, 96.5%). In A4, 85.4% (17.5, 94.5%) of cerebral amyloidosis was attributable to ε3 and ε4. The proportions of all-cause dementia attributable to ε3 and ε4 in UKB and FinnGen were 44.4% (95% CI: 18.2%, 62.2%) and 45.6% (30.6%, 56.9%), respectively. Without strong underlying risks from *APOE* ε3 and ε4, almost all AD and half of all dementia would not occur. Intervening on apolipoprotein E should be prioritised to facilitate dementia prevention.

## 1. Introduction

Strong associations between isoforms of apolipoprotein E (apoE) and risk of late-onset AD were established in the early 1990s.[1, 2] Subsequently, attention to apoE’s role in AD aetiology has been modest in comparison to research to understand and intervene upon AD’s pathophysiology – particularly the aggregation of cerebral amyloidosis and tauopathy.[3] However, only limited efficacy has been suggested by recent anti-amyloid therapeutic trials.[4, 5] The clinical benefits of modifying the neuropathological hallmarks of AD are still in doubt and a diversification of potential disease-modifying targets being addressed by AD researchers is warranted.[6–8] In this light, quantification of apoE’s contribution to AD is imperative because a causal role of variation in apoE isoforms on AD risk and progression is clear,[9] and a large proportion of dementia cases might be preventable by interventions related to this single molecule.

Common variation in the gene *APOE* produces three major isoforms of apoE in humans: ε2, ε3 and ε4. Relative to carriage of the allele encoding ε3 – the most common isoform with around 95% prevalence worldwide[10] – AD risk is much higher with ε4 carriage (∼28% prevalence) and much lower with ε2 carriage (∼14% prevalence).[11] The proportion of AD attributable to the detrimental ε4 isoform has been estimated in many settings, with population attributable fractions (PAFs) for this burden ranging considerably, from less than 20% to approximately 70%.[12–15] However, many of these estimates are substantially biased due to aspects of study design or calculations.[12, 14] Estimates for ε4 carriage also do not reflect the proportion of AD cases attributable to ε3, which is commonly misperceived as neutral for AD risk, despite ε3 substantially increasing AD risk relative to ε2 carriage.[14, 16]

By analysing data from approximately 470,000 participants across four independent studies, we provide estimates of the proportions of AD and all-cause dementia attributable to ε3 and ε4 carriage – i.e. the fraction of cases due to the combined impact of risk alleles inherited by most individuals. We also estimate the separate contributions of each allele to AD.

## 2. Methods

### 2.1 Study overview

We analysed data from three large cohorts – UK Biobank (UKB), FinnGen, and the Anti-Amyloid Treatment in Asymptomatic Alzheimer disease (A4) Study – along with re-analysis of published statistics from a case-control study by the Alzheimer’s Disease Genetics Consortium (ADGC). In each analysis, we modelled associations of *APOE* genotypes with AD ascertained by different methods (clinical diagnosis, neuropathology, and a combination of both), and calculated PAFs to indicate the burden of the outcomes attributable to *APOE* genotypes. We conducted analyses in several resources to provide a comprehensive estimate of view of apoE’s impact on AD burden. Clinically diagnosed AD without neuropathological confirmation is commonly misclassified (differential diagnoses with other causes of dementia being a major issue) and the presence of AD neuropathology *in vivo* does not always mean individuals will develop dementia.[17, 18] In UKB and FinnGen, we also produced equivalent statistics for all-cause dementia. Finally, we analysed published summary statistics from two genome-wide association studies (GWAS) to provide context for our results.

### 2.2 Samples

UKB is a multi-centre cohort study that recruited approximately 502,000 participants aged 39–73 years at assessment sites in England, Scotland, and Wales between 2006 and 2010.[19] Here, we used data from participants who were aged at least 60 years at baseline, so that individuals had an appreciable risk of late-onset AD within current follow-up. The sample was restricted to those with genotypic data, after exclusions for failing sample-level genetic quality control (genetic/phenotypic sex mismatches, excess heterozygosity, aneuploidy), the random removal of one individual from related pairs, and individuals who may have had ε3r alleles of *APOE* (n*=*171,105).

FinnGen is a public-private research project aggregating genotypic data from newly and historically collected samples in numerous Finnish biobanks, along with disease ascertainment via electronic health record (EHR) linkage.[20] The methods of the FinnGen study, including those for genotyping and endpoint definitions, are described in detail by Kurki et al.[20] We analysed individual-level data in the 13^th^ data release for individuals who were aged at least 60 years at first record of AD/dementia (for cases) or death / end of follow-up (for controls). The total analytical sample was 289,150 for the AD analysis.

The A4 Study is a multi-national randomised clinical trial that recruited eligible participants aged 65 to 85 years in the US, Canada, Australia and Japan between 2014 and 2017.[21] It implemented a 240-week intervention of the anti-amyloid therapy solanezumab to individuals with presymptomatic AD, following baseline screening of cognitive function and the presence of cerebral amyloidosis assessed by amyloid positron emission tomography (PET) scans. Here, we use data from participants who had baseline PET scans prior to the intervention phase of the study. After exclusions for missing data on *APOE* genotypes and self-reported ethnicity, the analytical sample was 4,415.

The ADGC has previously published associations of *APOE* genotypes with AD risk from a case-control study of individuals with and without confirmation of AD neuropathology at autopsy.[22] The primary sample in this analysis included 4018 deceased AD cases meeting the clinical and neuropathological criteria for AD, along with 989 deceased controls without cognitive impairment at their last clinical assessment in the study prior to death and without meeting the neuropathological criteria of AD postmortem (total N=5,007). For a comparison of results derived from the ADGC’s primary analytical sample, we also made use of a secondary analysis by the ADGC of a sample without postmortem neuropathological assessments, which included 10,430 cases meeting clinical criteria for probable AD and 13,427 cognitively normal controls.

### 2.3 APOE coding

In all samples, *APOE* ε2/ε3/ε4 alleles were coded from single nucleotide polymorphisms rs7412 and rs429358, measured by standalone genotyping or genotyped or hard-called imputed microarray data.[19–22] We analysed the relative risk of outcomes for each of the five risk-increasing genotypes (ε2/ε3, ε3/ε3, ε2/ε4, ε3/ε4, ε4/ε4) separately, relative to individuals with the lowest risk ε2/ε2 genotype. Individuals with an ε3/ε3 genotype are typically used as the reference group in analyses of *APOE* because ε3/ε3 is the most common genotype. However, this is inappropriate for calculating attributable fractions, where individuals with the lowest risk from the exposure of interest should be used as the reference category[23, 24] – those of ε2/ε2 genotype in this instance. This allows the total disease burden attributable to exposure to all risk-increasing genotypes to be calculated. The *APOE* variants were in Hardy-Weinberg equilibrium in all studies.

### 2.4 Ascertainment of outcomes

In UKB, AD was ascertained through linked electronic health records (EHR) and death records available up to July 2024 (minimum/maximum follow-up: 13.8 / 18.3 years). All-cause dementia was identified from a combination of self-report of ‘dementia or Alzheimer’s disease or cognitive impairment’ at baseline and record-linked follow-up. EHR-based ascertainment of both outcomes included Hospital Episode Statistics and death records for the full cohort using the cohort’s algorithmically defined outcomes.[25, 26] Diagnostic codes from primary care records were also used for the ∼45% of the cohort for which linkage to records from general practices had been arranged at the time of analysis (primary care codes are listed in Supplemental Table 1).

In FinnGen data release 13, ascertainment of AD and all-cause dementia was based on standardised endpoints. The definitions are based on combinations of hospital discharge, cause of death, and drug purchase reimbursement records from 1964 (for drug reimbursements) or 1969 (for hospital discharge and cause of death records) until November 2024. Codes are listed in Supplemental Tables 2 and 3.

In A4, cerebral amyloidosis was ascertained from amyloid-β positron emission tomography (PET) scans at the study’s baseline assessments prior to intervention.[21] As recommended to us by the A4 Study coordinators, PET standardised uptake value ratio (SUVr) values above a threshold of ≥1.15 indicated a positive amyloid-β scan (Aβ+).

In ADGC, AD neuropathology was scored according to the CERAD 4-point scale for neuritic Aβ plaque severity and the Braak 0-VI staging criteria for neurofibrillary tangle burden.[22] Cases and controls had been assessed for dementia prior to death according to DSM-IV or NINCDS/ADRDA criteria, and where available, Clinical Diagnostic Ratings (CDR). Assessments are described in further detail by Reiman et al.[22]

### 2.5 Statistical analysis

In UKB, risks of AD and all-cause dementia by *APOE* genotype were estimated using generalised linear models with a binomial distribution and log link (log-binomial modelling), which yield risk ratios (RR). Log-binomial models were adopted for analyses rather than survival analysis because of the sample’s mix of prevalent and incident cases, and because RRs are used in the formula for PAFs. We were unable to implement log-binomial models in A4 (non-convergence due to estimated probabilities of the outcome for some ε4/ε4 individuals being close to one) or FinnGen (log-binomial modelling is not currently an option in the software package REGENIE, which was necessary to implement to account for relatedness with linear mixed models in association testing in biobank-scale data). Instead, in these datasets, logistic regression was used to model associations and odds ratios (ORs) were converted to RRs for use in the PAF calculations, as were ORs in the ADGC data (detailed in the Supplemental Information).[27]

In UKB, FinnGen and A4 analyses, regression models were adjusted for age, sex and other covariates as follows. UKB and FinnGen additionally included adjustment for the first ten principal components derived from genome-wide microarray data (A4 lacked microarray data necessary to derive components on the full analytical sample), and FinnGen models also adjusted for genotyping batch. UKB and A4 also included adjustment for self-reported ethnicity (the FinnGen sample included participants only of Finnish ancestry). In UKB, ethnicity was entered as a binary variable for White/Other due to small numbers of ethnic minorities being present in some *APOE* genotype groups. In A4, a four-category variable was created for ‘White’, ‘Black’, ‘Asian’ or ‘Other’ (including those of mixed ancestry), with dummy variables comparing each group to those of White ethnicity entered in models (due to White being the category with the largest sample size). UKB models also adjusted for genotyping array type. FinnGen analyses accounted for relatedness among participants by using mixed modelling in the REGENIE software (v3.2.6).[28] The FinnGen REGENIE analysis was performed in dominant test mode on custom genotype data where *APOE* ε2/ε2 was encoded as the reference allele for the other genotype groups as alternate alleles.

We calculated PAFs based on case fractions across exposed groups and RRs per exposure group, an approach which can be used for multi-level exposures (formula ‘2C’ in [23]). 95% confidence intervals for PAFs were derived using the lower and upper confidence intervals for RRs. PAFs and their confidence intervals were converted from fractions to percentages.

In ADGC data, we also estimated the separate contributions of ε3 and ε4 to the total PAF for the outcomes from the sums of PAFs from individual genotypes including each allele separately (e,g, ε2/ε3 and ε3/ε3 for ε3 carriage) along with the estimated shares of each allele to the PAF due to ε3/ε4 (see Supplemental Information for further details).

To provide context for the magnitude of the PAF of AD and dementia due to ε3 and ε4, we also calculated PAFs for AD attributable to other genomic loci besides *APOE*. We used data from one of the largest genome-wide association studies (GWAS) of AD to date to identify hits within the top nine loci beyond *APOE* ranked by size of PAF for AD.[29] For a wider comparison of the magnitude of PAFs for AD to PAFs for loci related to other common diseases, we calculated the equivalent statistics for coronary artery disease (CAD) using summary statistics from one of the largest CAD GWAS to date.[30] We used the same approach for AD risk loci to calculate PAFs for the top hits at 65 then-known loci for CAD, and ranked these to use the 10 largest PAFs in our results. Calculations are described in the Supplemental Information. It is worth noting that various combinations of PAFs for these genetic loci can be summed to more than 100%. In multifactorial diseases without mutually exclusive causes, PAFs can overlap.[31, 32]

### 2.6 Ethics

All study participants had given informed consent and each study had received ethical / institutional review board approvals, as described previously.[19–22] An extended ethics statement for FinnGen is included in the Supplemental Information.

### 2.7 Patient and Public Involvement

This research involved secondary analysis of existing datasets and we did not involve patients or members of the public in the planning, conduct or interpretation of the study.

## 3. Results

Sample characteristics are displayed in Table 1. As expected, carriage of either ε3 or ε4 was highly prevalent in the full samples of UKB (99.4%), FinnGen (99.7%) and A4 (99.4%), and higher in AD cases (99.9%) than controls (98.1%) in the ADGC.

**Table 1.**
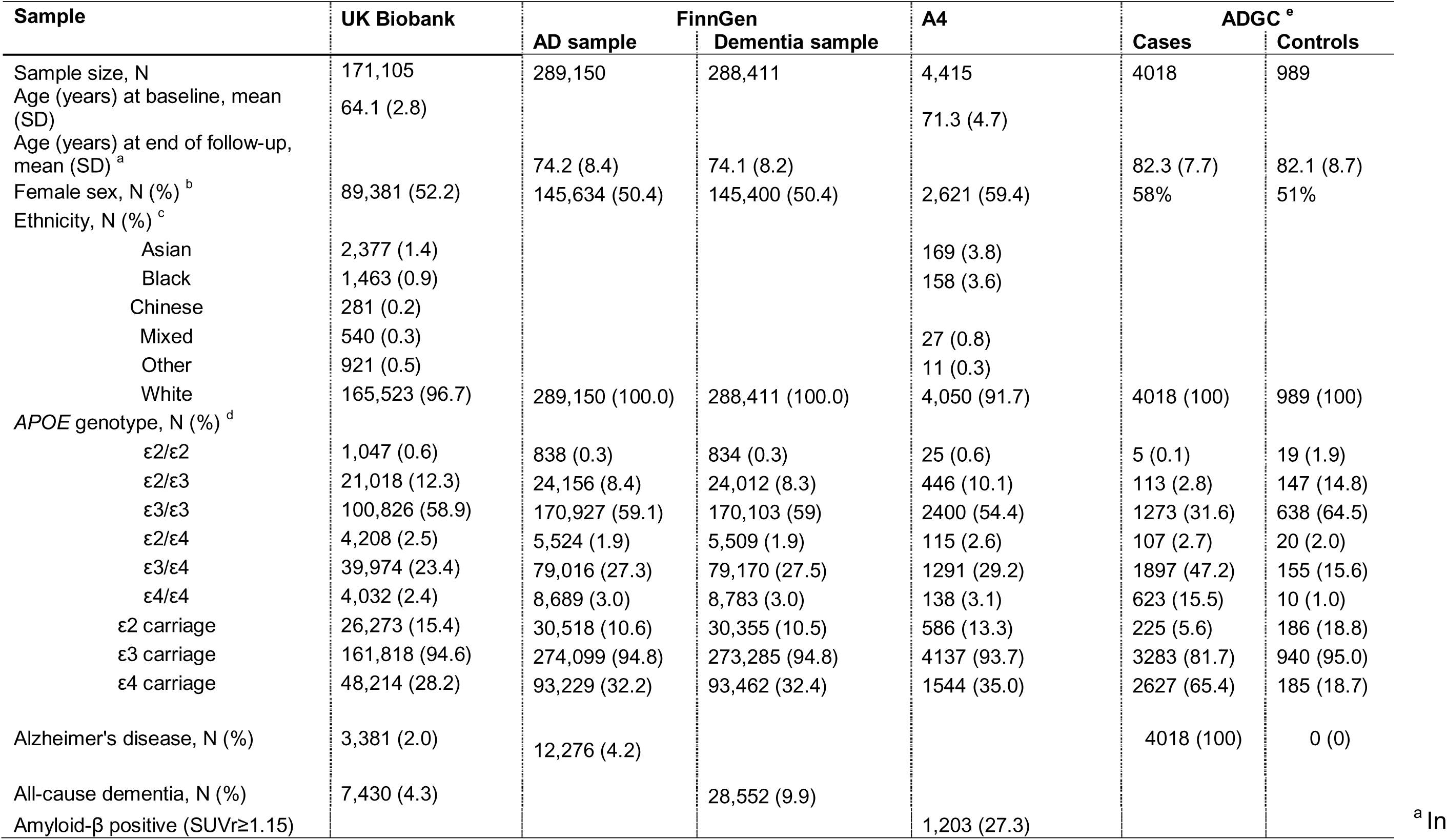
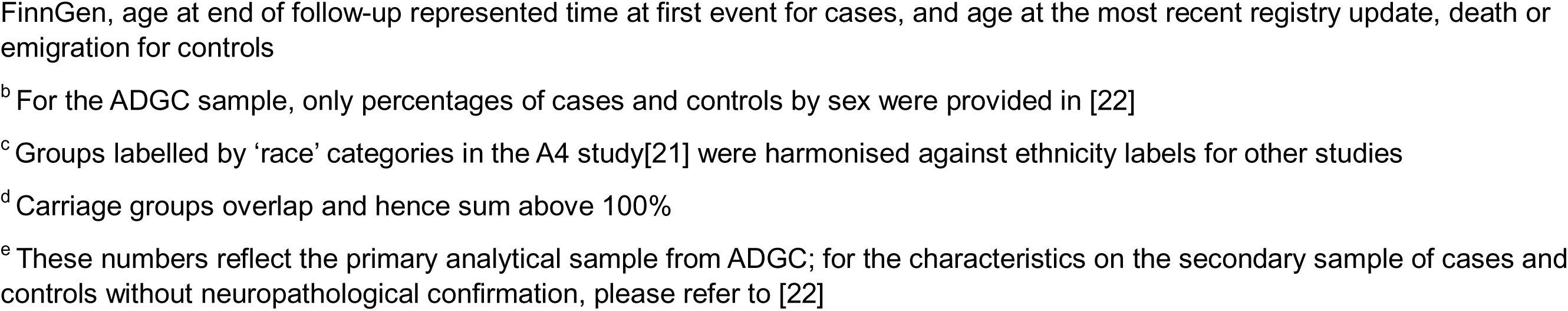
Characteristics of the analytical samples.

| Sample | UK Biobank | FinnGen |  | A4 | ADGC<sup>e</sup> |  |
| --- | --- | --- | --- | --- | --- | --- |
|  |  | AD sample | Dementia sample |  | Cases | Controls |
| Sample size, N | 171,105 | 289,150 | 288,411 | 4,415 | 4018 | 989 |
| Age (years) at baseline, mean (SD) | 64.1 (2.8) |  |  | 71.3 (4.7) |  |  |
| Age (years) at end of follow-up, mean (SD)<sup>a</sup> |  | 74.2 (8.4) | 74.1 (8.2) |  | 82.3 (7.7) | 82.1 (8.7) |
| Female sex, N (%)<sup>b</sup> | 89,381 (52.2) | 145,634 (50.4) | 145,400 (50.4) | 2,621 (59.4) | 58% | 51% |
| Ethnicity, N (%)<sup>c</sup> |  |  |  |  |  |  |
| Asian | 2,377 (1.4) |  |  | 169 (3.8) |  |  |
| Black | 1,463 (0.9) |  |  | 158 (3.6) |  |  |
| Chinese | 281 (0.2) |  |  |  |  |  |
| Mixed | 540 (0.3) |  |  | 27 (0.8) |  |  |
| Other | 921 (0.5) |  |  | 11 (0.3) |  |  |
| White | 165,523 (96.7) | 289,150 (100.0) | 288,411 (100.0) | 4,050 (91.7) | 4018 (100) | 989 (100) |
| APOE genotype, N (%)<sup>d</sup> |  |  |  |  |  |  |
| ε2/ε2 | 1,047 (0.6) | 838 (0.3) | 834 (0.3) | 25 (0.6) | 5 (0.1) | 19 (1.9) |
| ε2/ε3 | 21,018 (12.3) | 24,156 (8.4) | 24,012 (8.3) | 446 (10.1) | 113 (2.8) | 147 (14.8) |
| ε3/ε3 | 100,826 (58.9) | 170,927 (59.1) | 170,103 (59) | 2400 (54.4) | 1273 (31.6) | 638 (64.5) |
| ε2/ε4 | 4,208 (2.5) | 5,524 (1.9) | 5,509 (1.9) | 115 (2.6) | 107 (2.7) | 20 (2.0) |
| ε3/ε4 | 39,974 (23.4) | 79,016 (27.3) | 79,170 (27.5) | 1291 (29.2) | 1897 (47.2) | 155 (15.6) |
| ε4/ε4 | 4,032 (2.4) | 8,689 (3.0) | 8,783 (3.0) | 138 (3.1) | 623 (15.5) | 10 (1.0) |
| ε2 carriage | 26,273 (15.4) | 30,518 (10.6) | 30,355 (10.5) | 586 (13.3) | 225 (5.6) | 186 (18.8) |
| ε3 carriage | 161,818 (94.6) | 274,099 (94.8) | 273,285 (94.8) | 4137 (93.7) | 3283 (81.7) | 940 (95.0) |
| ε4 carriage | 48,214 (28.2) | 93,229 (32.2) | 93,462 (32.4) | 1544 (35.0) | 2627 (65.4) | 185 (18.7) |
| Alzheimer's disease, N (%) | 3,381 (2.0) | 12,276 (4.2) |  |  | 4018 (100) | 0 (0) |
| All-cause dementia, N (%) | 7,430 (4.3) |  | 28,552 (9.9) |  |  |  |
| Amyloid-β positive (SUVR≥1.15) |  |  |  | 1,203 (27.3) |  |  |

<sup>a</sup> In
FinnGen, age at end of follow-up represented time at first event for cases, and age at the most recent registry update, death or emigration for controls
<sup>b</sup> For the ADGC sample, only percentages of cases and controls by sex were provided in [22]
<sup>c</sup> Groups labelled by 'race' categories in the A4 study[21] were harmonised against ethnicity labels for other studies
<sup>d</sup> Carriage groups overlap and hence sum above 100%
<sup>e</sup> These numbers reflect the primary analytical sample from ADGC; for the characteristics on the secondary sample of cases and controls without neuropathological confirmation, please refer to [22]

In UKB, with reference to ε2/ε2 homozygotes, risk of both AD and all-cause dementia was increasingly higher in an expected gradient across genotypes ε2/ε3, ε3/ε3, ε2/ε4, ε3/ε4 and ε4/ε4 (Table 2). Results for all-cause dementia are presented in full in Supplemental Table 4. Summing the PAFs across all five risk-increasing genotypes, ε3 and ε4 together accounted for 75.7% (41.7, 89.8%) and 44.4% (18.2, 62.2%) of AD and all-cause dementia, respectively.

**Table 2:** Associations and proportions of outcomes attributable to *APOE* genotypes in each sample.

| Exposure | UKB (outcome: AD) | | FinnGen (outcome: AD) | | A4 (outcome: Aβ+) | | ADGC (outcome: neuropath. confirmed AD) | |
| --- | --- | --- | --- | --- | --- | --- | --- | --- |
|  | RR (95% CI) | PAF (95% CI), % | RR (95% CI) | PAF (95% CI), % | RR (95% CI) | PAF (95% CI), % | RR (95% CI) | PAF (95% CI), % |
| $\epsilon 2/\epsilon 2$ | Ref. | | Ref. | | Ref. | | Ref. | |
| $\epsilon 2/\epsilon 3$ | 1.68 (0.69, 4.07) | 2.0 (-2.2, 3.8) | 1.63 (1.00, 2.63) | 1.6 (0.0, 2.6) | 3.30 (0.48, 13.21) | 3.5 (-5.1, 4.6) | 2.94 (1.38, 5.71) | 1.9 (0.8, 2.3) |
| $\epsilon 3/\epsilon 3$ | 2.19 (0.91, 5.25) | 17.0 (-3.1, 25.4) | 2.29 (1.40, 3.71) | 22.7 (11.5, 29.4) | 4.27 (0.64, 14.88) | 25.9 (-16.5, 31.5) | 7.21 (2.73, 16.81) | 27.3 (20.1, 29.8) |
| $\epsilon 2/\epsilon 4$ | 4.03 (1.64, 9.91) | 1.8 (0.9, 2.2) | 4.44 (2.87, 6.79) | 1.9 (1.6, 2.1) | 8.36 (1.42, 19.58) | 2.7 (1.0, 2.9) | 17.23 (4.42, 46.83) | 2.5 (2.1, 2.6) |
| $\epsilon 3/\epsilon 4$ | 8.39 (3.50, 20.13) | 41.9 (34.0, 45.2) | 6.37 (4.32, 9.25) | 35.6 (32.5, 37.7) | 12.39 (2.80, 21.89) | 44.9 (32.0, 46.6) | 32.26 (12.48, 59.95) | 45.7 (43.4, 46.4) |
| $\epsilon 4/\epsilon 4$ | 23.36 (9.71, 56.19) | 12.9 (12.1, 13.2) | 9.79 (7.44, 12.73) | 9.7 (9.4, 10.0) | 20.32 (8.67, 24.24) | 8.2 (7.7, 8.3) | 70.81 (31.76, 92.23) | 15.3 (15.0, 15.3) |
| Total |  | 75.7 (41.7, 89.8) |  | 71.5 (54.9, 81.7) |  | 85.1 (19.2, 93.9) |  | 92.7 (81.4, 96.5) |

*PAF – population attributable fraction; RR – risk ratio*

In FinnGen, the same gradient and similar magnitudes of associations of risk of AD and all-cause dementia was observed across *APOE* genotypes as in UKB, though the association of ε4/ε4 with AD was notably more modest (Table 2; Supplemental Table 4). The corresponding PAFs were 71.5% (54.9, 81.7%) for AD and 45.6% (30.6, 56.9%) for all-cause dementia.

In A4, cerebral amyloidosis was present in only one of 25 (4.0%) ε2 homozygotes in the sample, and in 1202 of 4390 (27.4%) of ε3 and ε4 carriers. The same gradient of relative risk across *APOE* genotypes was present for amyloidoisis as was seen for AD risk in UKB and FinnGen (Table 2; Supplemental Table 4). The PAF for cerebral amyloidosis attributable to ε3 and ε4 was 85.1%; (95% CI: 19.2%, 93.9%). It is also worth noting from Supplemental Figure 1 that the sole ε2 homozygote designated as Aβ+ had a lower SUVr than the means for ε3/ε3 and ε4/ε4 Aβ+ individuals. In other words, even where an ε2 homozygote had evidence of amyloidosis, this individual was positioned lower than individuals with other *APOE* genotypes on a continuum of the degree of pathology being detected.

In ADGC data, after re-orienting ORs to be with reference to ε2/ε2 individuals (shown in Supplemental Table 5), a very strong increasing gradient of RRs across *APOE* genotypes ε2/ε3, ε3/ε3, ε2/ε4, ε3/ε4 and ε4/ε4 2 was observed for risk of neuropathologically confirmed AD (Table 2, Supplemental Table 4). This equated to a PAF of 92.7% (95% CI: 81.4%, 96.5%) attributable to ε3 and ε4 together in our primary analysis, where RRs were generated from ORs assuming a 1% baseline probability of developing AD in ε2/ε2 individuals. In extended calculations that partitioned this PAF into the separate contributions of the ε3 and ε4 alleles (Supplemental Table 6), ε4 was estimated to account for 56.9% of neuropathologically confirmed AD (95% CI: 36.0%, 63.0%), with the remainder specifically attributable to ε3 (35.8%; 95% CI: 22.1%, 58.3%). In secondary analyses, the PAF for AD in the ADGC sample where neuropathological assessments of cases and controls were not available was lower (69.8%, 95% CI: 47.7%, 82.2%), and similar in magnitude to PAF estimates from UKB and FinnGen (Supplemental Table 4). Sensitivity analyses varying the assumption of baseline probability of AD from 1% to 5% among ε2 homozygotes did not markedly affect these PAF estimates (Supplemental Table 7).

Finally, comparing the total PAF for ε3 and ε4 carriage from the ADGC analysis to other genetic risk loci for AD, and to risk loci for CAD, no other PAF for loci for either disease exceeded 36% (Figure 1). Calculations are shown in Supplemental Tables 8 and 9.

**Figure 1:**
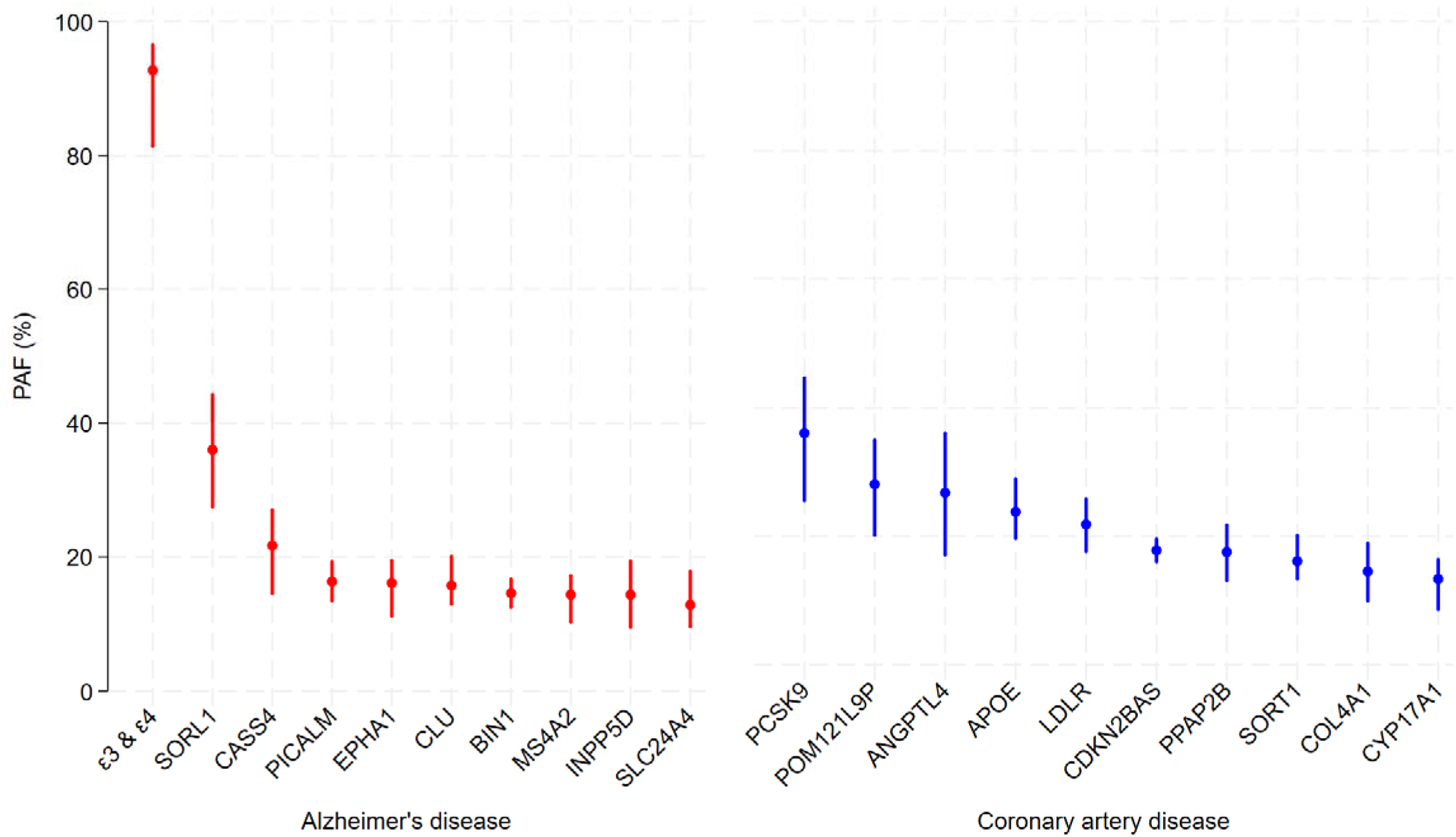
Proportions of AD and CAD that could be prevented by targeting molecular pathways related to the top ten genetic risk loci for each disease, ranked by size of PAF *PAF – population attributable fraction* Footnotes : ^1^ The *APOE* ε3 and ε4 estimate for AD is based on our PAF estimate from ADGC data, alongside statistics for other risk loci from GWAS. In the CAD GWAS, the PAF estimate for the *APOE* locus estimate was based solely on the odds of CAD for risk-conferring allele (C) of genotype rs7412, i.e. this estimate reflects the PAF for genotypes ε3/ε3, ε3/e4 and ε4/ε4 relative to ε2/ε2, ε2/ε3 and ε2/ε4 carriers (hence it may underestimate the contribution of *APOE* genotypes to CAD). ^2^ Note that the confidence intervals for the PAF of AD attributable to ε3 and ε4 are not symmetric because PAFs plateau as they approach 100%.

## 4. Discussion

### 4.1 Summary

Our findings, spanning several large population-based and case-control datasets, consistently indicate that if interventions could eliminate the detrimental effects of ε3 and ε4 carriage, we could expect to prevent most AD and a large proportion of all dementia. In other words, the two common risk alleles in *APOE* are major component causes for AD, and without them most disease would not occur. The existence of the ε3 allele is specifically contributing to much AD burden, in addition to the fraction of disease attributable to ε4.

### 4.2 Study-level interpretation

Some aspects of individual analyses warrant attention. First, in UKB and FinnGen, associations of *APOE* genotypes with AD risk were more modest than may have been anticipated, considering magnitudes observed in other settings.[11, 29] This may have been due to several sources of bias in outcome ascertainment: limited record linkage (not yet fully extending to primary care or mental health service records)[25], limitations to the use of clinical codes, no follow-up with cognitive assessments of the whole surviving cohort, and incomplete measurement of outcome lifetime risks – e.g. the youngest participants in the UKB sample were aged approximately 74 years at the end of current follow-up. Second, in A4, the contribution of ε4/ε4 to the burden of amyloidosis among the sample was lower than anticipated. This may have resulted from selection effects in the A4 Study’s design, where ε4/ε4 participants at higher risk of cognitive impairment and established amyloidosis may have been less likely to volunteer for the study and to pass initial screening for cognitively normal individuals to bring forward for amyloid PET scans.[21] This potential downward bias for ε4/ε4 individuals is reflected in the distribution of SUVr values by genotype (Supplemental Figure 1), with the mean values of ε2, ε3 and ε4 homozygotes above the Aβ+ threshold denoted. Aβ+ ε4 homozygotes had similar mean SUVr values to Aβ+ ε3 homozygotes (1.37 and 1.33, respectively). Third, in comparing primary and secondary analyses in ADGC data (with and without neuropathological assessment for cases and controls, respectively), less pronounced RRs and PAFs for the secondary analysis could indicate the influence of misclassification / quality of differential diagnosis among dementia cases in samples where neuropathology was not assessed. Finally, as our analysis of GWAS data for AD and CAD indicates, the preponderance of AD burden attributable to genetic variation at a single locus may be exceptional among common, complex chronic diseases. We also draw attention to the fact that PAFs for different loci can overlap.[31] These circumstances can imply that there is some degree of interaction between the molecular entities that each gene encodes – as is well established in the case of *LDLR* and *PCSK9* in relation to regulation of circulating low-density lipoprotein cholesterol and, hence, CAD risk.[33] Another possibility explaining overlapping PAFs is that these molecular pathways could be distinct but converge on a common mediating factor in a disease, e.g. separate pathways each promoting the aggregation of AD neuropathology.

### 4.3 Interpretation in relation to other research

Some previous research and commentaries have implied that most AD can be attributed to the risk incurred by the ε3 and ε4 alleles of *APOE*, albeit without having modelled risk relative to ε2/ε2 individuals directly, as we have.[2, 12, 34, 35] Most previous PAF estimates have addressed the contribution of the ε4 allele alone to AD and all-cause dementia in different settings.[13, 14, 36] These have generally been flawed for several reasons including biases in study design,[12] and errors in calculations have occurred when allele frequency is mistaken for allele prevalence.[14, 37] A major issue is that estimates commonly combine ε3 and ε2 carriers in a reference group, which means risk of AD due to ε4 carriage is compared to risk with carriage of both moderate and low-risk alleles. The current analysis differs by estimating these fractions directly in data with a large number of ε2 homozygotes present, by combining different outcomes including clinical diagnoses and AD neuropathological assessment, and by partitioning estimates of disease burden attributable to the ε3 and ε4 alleles separately. Very large analytical samples are required to use rare ε2 homozygotes as the reference group in analyses, and there has been little recognition among dementia researchers that the ε3 allele should also be considered risk-increasing for AD. Nonetheless, we estimate that the ε3 allele alone could be responsible for a third or more of AD due to ε3 conferring considerable risk to most individuals.

We emphasise that our results are not suggesting that AD is monogenic. These results should be interpreted in line with theories for complex disease aetiology, which allow for multiple contributing causes to aggregate for causation [31, 32, 38]. AD is clearly multifactorial, but its other component causes beyond apoE’s role are of much less consequence without the underlying risk from ε3 or ε4 carriage that most people inherit. Put differently, if all individuals inherited an ε2/ε2 genotype, most AD would not occur, regardless of what other risk factors people experience. It should also be noted that PAFs are conceptually distinct from heritability, and that heritability analyses are not informative for assessing disease burden attributable to specific causes.[31]

### 4.4 Strengths and limitations

Strengths of this study are the use of samples large enough to conduct models with relatively rare ε2 homozygotes as the reference group, and comparisons of findings from multiple data sources with different designs, sample characteristics and outcome definitions, all with consistent findings. A limitation is that our PAF estimates were somewhat imprecise (and findings from A4 particularly so) due to measures of disease risk among a relatively rare reference group, where chance differences in case prevalence/incidence would be more influential. However, point estimates from all analyses for AD were consistently large and yielded similar conclusions. Second, our analytical samples consisted predominantly or exclusively of individuals of European ancestry. *APOE* associations with AD risk differ by ancestry,[11] and hence these results may not be generalisable to other ethnic groups. Further research should estimate PAFs in samples of non-European ancestry according to the approach that we took. Third, PAF estimates are sensitive to aspects of study design, such as follow-up periods, the extent of outcome ascertainment, selection biases (including differential bias across *APOE* genotypes due to cardiovascular morbidity and mortality).[30],[12] It is therefore reassuring that our findings and interpretation are not materially different across different samples, study designs and definitions of AD. Finally, attributable fractions assume that the exposure of interest is a cause of the disease being investigated and not based on biased estimates of association, which can arise, for example, due to confounding or reverse causation.[39] This may often be an unrealistic assumption for environmental and clinical risk factors that have been investigated for AD and all-cause dementia.[40] However, due to the properties of genetic inheritance, risk estimates for genetic variants such as the *APOE* alleles are not subject to reverse causation and are unlikely to be affected by confounding.[41] Genetic and experimental evidence also imply that the effects of variation in *APOE* on AD and all-cause dementia risk are highly unlikely to be due to nearby co-inherited genetic variation (in linkage disequilibrium with *APOE*), rather than the *APOE* variants per se.[9] Hence, *APOE* variants unequivocally cause AD and PAFs for these variants provide robust estimates of disease burden attributable to the variation in question.

### 4.5 Conclusions

Considering that most AD could be prevented (or at least delayed) by reducing the risk conferred by apoE, understanding the protein’s detrimental effects should be given proportionate research attention and funding. These should include efforts to understand the distinct functional properties of the ε3 isoform responsible for AD risk, relative to properties of the ε2 isoform and other lower-risk variants.[42, 43] There is considerable scope to target apoE with interventions. With gene editing, transfer and silencing approaches, genetic risk is now directly modifiable. Moreover, many strategies exist to target apoE at the protein level or its interactions with molecular intermediaries including immunotherapy and small molecule structural correctors.[44, 45] However, only one therapy targeting apoE (LX1001)[46] is currently being trialled for AD in humans – less than 1% of potential therapies in registered trials.[47] A rebalancing of therapeutic development for AD (as well as basic research) towards apoE is warranted. Prioritising direct research into apoE does not preclude investigations into other genetic or environmental factors that could be mediating or modifying the effects of apoE on AD, or research into factors that may be distinct component causes of these outcomes (both scenarios include research addressing cerebral amyloidosis and tauopathy). Nonetheless, establishing how, when, and in which cell types apoE influences AD risk – and how its deleterious effects can be mitigated – is clearly paramount to AD prevention and treatment.

## Data Availability

All UKB data used in this research are available to researchers who register with UK Biobank and request access to them as part of an approved project: https://www.ukbiobank.ac.uk/. A4 Study data are made publicly available to researchers via the Imaging and Data Archive (https://ida.loni.usc.edu/). Individual-level data held by FinnGen is accessible to qualifying researchers in sponsoring institutions upon approval of a project application by the study's scientific committee.

https://www.a4studydata.org/

https://www.finngen.fi/en

## Abbreviations

A4: Anti-Amyloid Treatment in Asymptomatic Alzheimer disease study
ADGC: Alzheimer’s Disease Genetics Consortium
ApoE: Apolipoprotein E
CAD: Coronary artery disease
CI: Confidence interval
EHR: Electronic Health Records
OR: Odds ratio
PAF: Population attributable fraction
RR: Risk ratio
UKB: UK Biobank
Aβ+: Amyloid-β positive

**Acknowledgements**

*UK Biobank*

This research has been conducted using the UK Biobank Resource under application number 71702. This work uses data provided by patients and collected by the NHS as part of their care and support. Copyright © 2023, NHS England. Re-used with the permission of the NHS England and UK Biobank. All rights reserved.

*FinnGen*

We want to acknowledge the participants and investigators of the FinnGen study. The FinnGen project is funded by two grants from Business Finland (HUS 4685/31/2016 and UH 4386/31/2016) and the following industry partners: AbbVie Inc., AstraZeneca UK Ltd, Biogen MA Inc., Bristol Myers Squibb Inc. (and Celgene Corporation & Celgene International II Sàrl), Genentech Inc., Merck Sharp & Dohme LCC, Pfizer Inc., GlaxoSmithKline Intellectual Property Development Ltd., Sanofi US Services Inc., Maze Therapeutics Inc., Johnson&Johnson Innovative Medicine Inc., Novartis AG, Boehringer Ingelheim International GmbH and Bayer AG. Following biobanks are acknowledged for delivering biobank samples to FinnGen: Auria Biobank (www.auria.fi/biopankki), THL Biobank (www.thl.fi/biobank), Helsinki Biobank (www.helsinginbiopankki.fi), Biobank Borealis of Northern Finland (https://www.ppshp.fi/Tutkimus-ja-opetus/Biopankki/Pages/Biobank-Borealis-briefly-in-English.aspx), Finnish Clinical Biobank Tampere (www.tays.fi/en-US/Research_and_development/Finnish_Clinical_Biobank_Tampere), Biobank of Eastern Finland (www.ita-suomenbiopankki.fi/en), Central Finland Biobank (www.ksshp.fi/fi-FI/Potilaalle/Biopankki), Finnish Red Cross Blood Service Biobank (www.veripalvelu.fi/verenluovutus/biopankkitoiminta), Terveystalo Biobank (www.terveystalo.com/fi/Yritystietoa/Terveystalo-Biopankki/Biopankki/) and Arctic Biobank (https://www.oulu.fi/en/university/faculties-and-units/faculty-medicine/northern-finland-birth-cohorts-and-arctic-biobank). All Finnish Biobanks are members of BBMRI.fi infrastructure (https://www.bbmri-eric.eu/national-nodes/finland/). Finnish Biobank Cooperative -FINBB (https://finbb.fi/) is the coordinator of BBMRI-ERIC operations in Finland. The Finnish biobank data can be accessed through the Fingenious^®^ services (https://site.fingenious.fi/en/) managed by FINBB.

*A4*

The A4 Study was a secondary prevention trial in preclinical Alzheimer’s disease, aiming to slow cognitive decline associated with brain amyloid accumulation in clinically normal older individuals. The A4 Study was funded by a public-private-philanthropic partnership, including funding from the National Institutes of Health-National Institute on Aging, Eli Lilly and Company, Alzheimer’s Association, Accelerating Medicines Partnership, GHR Foundation, an anonymous foundation, and additional private donors, with in-kind support from Avid Radiopharmaceuticals, Cogstate, Albert Einstein College of Medicine and the Foundation for Neurologic Diseases. The companion observational Longitudinal Evaluation of Amyloid Risk and Neurodegeneration (LEARN) Study was funded by the Alzheimer’s Association and GHR Foundation. The A4 and LEARN Studies were led by Dr. Reisa Sperling at Brigham and Women’s Hospital, Harvard Medical School, and Dr. Paul Aisen at the Alzheimer’s Therapeutic Research Institute (ATRI) at the University of Southern California. The A4 and LEARN Studies were coordinated by ATRI at the University of Southern California, and the data are made available under the auspices of Alzheimer’s Clinical Trial Consortium through the Global Research & Imaging Platform (GRIP). The complete A4 Study Team list is available on: https://www.actcinfo.org/a4-study-team-lists/. We would like to acknowledge the dedication of the study participants and their study partners who made the A4 and LEARN Studies possible.

## Data Availability

All UKB data used in this research are available to researchers who register with UK Biobank and request access to them as part of an approved project: https://www.ukbiobank.ac.uk/. A4 Study data are made publicly available to researchers via the Imaging and Data Archive (https://ida.loni.usc.edu/). Individual-level data held by FinnGen is accessible to qualifying researchers in sponsoring institutions upon approval of a project application by the study’s scientific committee. The data fields and script used in the analyses will be made available at the following site upon publication of this article: https://github.com/dylwil/ad_apoe_paf

