## Supplementary information and supplementary tables 1 to 3 for "The proportion of Alzheimer’s disease attributable to apolipoprotein E"

*Group authorship, constituent authors listed in the Supplemental Table 10.

**Supplemental Information**

*Conversion of ORs to RRs for FinnGen, A4 and ADGC results*

In FinnGen and A4, OR-to-RR conversion calculations used the baseline probabilities of the outcomes in each sample, i.e. the fraction of ε2 homozygotes with the outcome. ORs in the ADGC data were re-oriented to be with reference to ε2/ε2 individuals, rather than ε3/ε3, on the basis that differences in odds between the *APOE* genotype groups is additive on the log-scale. Given that the case-control study design of the ADGC yielded no direct measure of baseline probability of the outcome among the ε2/ε2 group, we converted ORs to RRs using a range of realistic baseline probabilities of AD based on lifetime risk estimates stratified by *APOE* genotype.^1^ We adopted 1% baseline risk for the conversion in our primary analysis, and repeated OR-to-RR conversions with baseline probabilities ranging up to 5% in sensitivity analyses.

*Process of deriving separate PAFs for ε3 and ε4*

For ADGC results, the specific PAF for ε3/ε4 was partitioned into ε4 and ε3 contributions according to the ratio by which ε2/ε4 to ε2/ε3 genotypes increase the risk of AD, i.e. according to the individual effects of the two alleles on AD risk. For instance, a ratio of 2:1 for the RRs of ε2/ε4 to ε2/ε3 for an outcome would assign 66.6% of the PAF for the ε3/ε4 genotype to ε4 specifically, with the remainder attributed to ε3. This assumes that any contribution to an interaction between the two alleles in relation to AD risk is proportionate to each allele’s effects on AD risk in isolation among ε2/ε4 and ε2/ε3 carriers. The construction of confidence intervals for the separate contributions of ε3 and ε4 was based on the following calculations:

- Lower CI limit for ε3 PAF = lower PAF estimate for ε2/ε3 + lower PAF estimate for ε3/ε3 + share of PAF estimate for ε3/ε4 based on ratio of upper RR for ε2/ε4 to lower RR for ε2/ε3
- Upper CI limit for ε3 PAF = upper PAF estimate for ε2/ε3 + upper PAF estimate for ε3/ε3 + share of PAF for ε3/ε4 based on ratio of lower RR for ε2/ε4 to upper RR for ε2/ε3
- Lower CI limit for ε4 PAF = lower PAF estimate for ε2/ε4 + lower PAF estimate for ε4/ε4 + share of PAF for ε3/ε4 based on ratio of lower RR for ε2/ε4 to upper RR for ε2/ε3
- Upper CI limit for ε4 PAF = upper PAF estimate for ε2/ε4 + upper PAF estimate for ε4/ε4 + share of PAF for ε3/ε4 based on ratio of upper RR for ε2/ε4 to lower RR for ε2/ε3

*PAF calculations from AD and CAD GWAS data*

PAF calculations for risk loci identified by GWAS of AD and CAD used an alternate formula based on estimated exposure prevalence in the underlying population rather than exposure distribution among case fractions, because we did not have case fractions per genotype for the GWAS results (as we did for *APOE* genotypes in our main analysis):

$AF (population) =\frac{{Prevalence}_{exposure}(RR-1)}{1+{Prevalence}_{exposure}(RR-1)}$	(1)

For the risk-increasing allele of the top hits in each of the 23 loci identified by the AD GWAS (21 confirmed, two suggestive)^2^ and 65 known risk loci for CAD,^3^ we first identified allele frequency *p* among 503 individuals of European ancestry in the 1000 Genomes project, phase 3.^4^ We used allele frequencies derived from these population-based reference data to derive exposure prevalence for each genotype rather than using the in-sample allele frequencies because allele frequencies from case-control data are skewed upward among cases (and downward among controls) for risk variants. We then calculated genotype frequencies for homozygous carriers of the risk allele (*p*^2^) and heterozygous carriers of the risk allele (2 × *p* x (1-*p*)). The GWAS estimated odds by assuming additive effects of variants, so we estimated overall PAFs for each variant as a sum of a PAF for homozygous carriers of the allele and a PAF for heterozygous carriers of the allele. The genotype frequencies of homozygotes and heterozygotes were entered as the prevalence of the exposure in equation (1). For homozygotes, odds ratios were recalculated as exp(2 × log-odds) reported by the GWAS to reflect them having double the risk in an additive model; for heterozygotes, odds ratios were based on the reported log-odds. We used meta-analysed stage 1 and stage 2 association statistics for these odds ratios. Given that these calculations used odds rather than risk ratios, the PAFs for AD and CAD risk loci besides *APOE* may have been slightly inflated. However, the deviation between odds and risk is minor for modest association magnitudes, and no odds ratio used in these calculations exceeded 2.08 (with all but one being ≤1.32).

**FinnGen Ethics statement and materials & methods:**

Study subjects in FinnGen provided informed consent for biobank research, based on the Finnish Biobank Act. Alternatively, separate research cohorts, collected prior the Finnish Biobank Act came into effect (in September 2013) and start of FinnGen (August 2017), were collected based on study-specific consents and later transferred to the Finnish biobanks after approval by Fimea (Finnish Medicines Agency), the National Supervisory Authority for Welfare and Health. Recruitment protocols followed the biobank protocols approved by Fimea. The Coordinating Ethics Committee of the Hospital District of Helsinki and Uusimaa (HUS) statement number for the FinnGen study is Nr HUS/990/2017.

The FinnGen study is approved by Finnish Institute for Health and Welfare (permit numbers: THL/2031/6.02.00/2017, THL/1101/5.05.00/2017, THL/341/6.02.00/2018, THL/2222/6.02.00/2018, THL/283/6.02.00/2019, THL/1721/5.05.00/2019 and THL/1524/5.05.00/2020), Digital and population data service agency (permit numbers: VRK43431/2017-3, VRK/6909/2018-3, VRK/4415/2019-3), the Social Insurance Institution (permit numbers: KELA 58/522/2017, KELA 131/522/2018, KELA 70/522/2019, KELA 98/522/2019, KELA 134/522/2019, KELA 138/522/2019, KELA 2/522/2020, KELA 16/522/2020), Findata permit numbers THL/2364/14.02/2020, THL/4055/14.06.00/2020, THL/3433/14.06.00/2020, THL/4432/14.06/2020, THL/5189/14.06/2020, THL/5894/14.06.00/2020, THL/6619/14.06.00/2020, THL/209/14.06.00/2021, THL/688/14.06.00/2021, THL/1284/14.06.00/2021, THL/1965/14.06.00/2021, THL/5546/14.02.00/2020, THL/2658/14.06.00/2021, THL/4235/14.06.00/2021, THL/4990/14.02.00/2023 Statistics Finland (permit numbers: TK-53-1041-17 and TK/143/07.03.00/2020 (earlier TK-53-90-20) TK/1735/07.03.00/2021, TK/3112/07.03.00/2021) and Finnish Registry for Kidney Diseases permission/extract from the meeting minutes on 4th July 2019.

The Biobank Access Decisions for FinnGen samples and data utilized in FinnGen Data Freeze 13 include: THL Biobank BB2017_55, BB2017_111, BB2018_19, BB_2018_34, BB_2018_67, BB2018_71, BB2019_7, BB2019_8, BB2019_26, BB2020_1, BB2021_65, BB22-0025-A01, BB22-0025-A03, BB23-0222-A01, BB22-0025-A04, BB22-0025-A06, BB22-0025-A08, THLBB2024_30. Finnish Red Cross Blood Service Biobank 7.12.2017, 13.11.2023, 001-2023, Helsinki Biobank HUS/359/2017, HUS/248/2020, HUS/430/2021 §28, §29, HUS/150/2022 §12, §13, §14, §15, §16, §17, §18, §23, §58, §59, HUS/128/2023 §18, BB22-0025-A01, BB22-0025-A02, BB22-0025-A05, BB22-0025-A07, BB22-0025-A09, BB22-0025-A10, BB22-0025-A03, BB23-0222-A01, BB22-0025-A04, BB22-0025-A06, BB22-0025-A08, Amendment_BB22-0025-A05, Decision allowing to continue data processing until 31st Aug 2027: BB_2021-0140, HUS/150/2022 §12, BB_2021-0139, HUS/150/2022 §13, BB_2021-0161,HUS/150/2022 §14, BB_2021-0164, HUS/150/2022 §15, BB_2021-0169, HUS/150/2022 §16, BB_2021-0170, HUS/150/2022 §17, BB_2021-0179, HUS/150/2022 §18, BB_2022-0262, HUS/150/2022 §58, BB22-0067, HUS/150/2022 §59, Auria Biobank AB17-5154 and amendment #1 (August 17 2020) and amendments BB_2021-0140, BB_2021-0156 (August 26 2021, Feb 2 2022), BB_2021-0169, BB_2021-0179, BB_2021-0161, AB20-5926 and amendment #1 (April 23 2020) and it´s modifications (Sep 22 2021), BB_2022-0262, BB_2022-0256, BB22-0025-A01, BB22-0025-A02, BB22-0025-A03, BB23-0222_A01, BB22-0025-A02, BB22-0025-A05, BB22-0025-A07, BB22-0025-A09, BB22-0025-A10, BB22-0025-A03, BB23-0222-A01, BB22-0025-A04, BB22-0025-A06, BB22-0025-A08, Decision allowing to continue data processing until 31st Aug 2027: AB20-5926, BB_2021- 0140, BB_2021-0156, BB_2021-0161, BB_2021- 0161, BB_2021-0164, BB_2021-0169, BB_2021-0179, BB_2022-0262, Biobank Borealis of Northern Finland_2017_1013, 2021_5010, 2021_5010 Amendment, 2021_5018, 2021_5018 Amendment, 2021_5015, 2021_5015 Amendment, 2021_5015 Amendment_2, 2021_5023, 2021_5023 Amendment, 2021_5023 Amendment_2, 2021_5017, 2021_5017 Amendment, 2022_6001, 2022_6001 Amendment, 2022_6006 Amendment, 2022_6006 Amendment_2, BB22-0067, 2022_0262, 2022_0262 Amendment, BB22-0025-A01, BB22-0025-A02, BB22-0025-A05, BB22-0025-A07, BB22-0025-A09, BB22-0025-A10, BB22-0025-A03, BB23-0222-A01, BB22-0025-A04, BB22-0025-A06, BB22-0025-A08, Decision allowing to continue data processing until 31st Aug 2027: BB/2021/5015, BB/2021/5017, BB/2021/5018, BB/2021/5023, BB/2022/6006, BB/2022/6001, BB/2022-0262, BB/2021/5010, Biobank of Eastern Finland 1186/2018 and amendment 22§/2020, 53§/2021, 13§/2022, 14§/2022, 15§/2022, 27§/2022, 28§/2022, 29§/2022, 33§/2022, 35§/2022, 36§/2022, 37§/2022, 39§/2022, 7§/2023, 32§/2023, 33§/2023, 34§/2023, 35§/2023, 36§/2023, 37§/2023, 38§/2023, 39§/2023, 40§/2023, 41§/2023, BB22-0025-A01, BB22-0025-A02, BB22-0025-A05, BB22-0025-A07, BB22-0025-A09, BB22-0025-A10, BB22-0025-A03, BB23-0222-A01, BB22-0025-A04, BB22-0025-A06, BB22-0025-A08, Decision allowing to continue data processing until 31st Aug 2027: MO-BB_2021-0179-A0, MO-BB_2021-0156_PRE-A01, BB_2021-0140, MO-BB_2021-0170_PRE-A0, MO-BB_2021-0169-A01, MO-BB_2022-0256-A01, MO-BB_2021-0161-A01, MO-BB_2021-0161-A02, BB22-0067-A01, MO-BB_2022-0262-A0, Finnish Clinical Biobank Tampere MH0004 and amendments (21.02.2020 & 06.10.2020), BB2021-0140 8§/2021, 9§/2021, §9/2022, §10/2022, §12/2022, 13§/2022, §20/2022, §21/2022, §22/2022, §23/2022, 28§/2022, 29§/2022, 30§/2022, 31§/2022, 32§/2022, 38§/2022, 40§/2022, 42§/2022, 1§/2023, BB2021-0140, BB22-0025-A01, BB_2021-0161, BB22-0025-A02, BB22-0025-A05, BB22-0025-A07, BB22-0025-A09, BB22-0025-A10, BB22-0025-A03, BB23-0222-A01, BB22-0025-A04, BB22-0025-A06, BB22-0025-A08, Decision allowing to continue data processing until 31st Aug 2027: BB_2021- 0140, BB_ 2021- 0161, BB_ 2021- 0179, BB_ 2021- 0156, BB_ 2021- 0169, BB_ 2021- 0170, BB22- 0067- A01, Central Finland Biobank 1-2017, BB_2021-0169, BB_2021-0179, BB_2022-0256, BB_2022-0262, Decision allowing to continue data processing until 31st Aug 2027 for projects: BB_2021-0179, BB22-0067,BB_2022-0262, BB_2021-0170, BB_2021-0164, BB_2021-0161, and BB_2021-0169, BB22-0025-A01, BB22-0025-A02, BB22-0025-A05, BB22-0025-A07, BB22-0025-A09, BB22-0025-A10, BB22-0025-A03, BB23-0222-A01, BB22-0025-A04, BB22-0025-A06, BB22-0025-A08, Terveystalo Biobank STB 2018001 and amendment 25th Aug 2020, Finnish Hematological Registry and Clinical Biobank decision 18th June 2021, Amendment 2nd January 2024 and Arctic biobank P0844: ARC_2021_1001, ARC_2023_3003 (BB22-0025-A01), BB22-0025-A03, BB23-0222-A01, BB22-0025-A04, BB22-0025-A06, BB22-0025-A08.

### **Supplemental Table 1: primary care diagnostic codes used to ascertain AD and all-cause dementia in UK Biobank**

| **Version** | **Code** | **Term** | **Code included in AD definition** |
| --- | --- | --- | --- |
| Read V3 (CTV3) | 1461. | H/O: dementia |  |
| Read V3 (CTV3) | A410. | Kuru |  |
| Read V3 (CTV3) | A411. | Creutzfeldt-Jakob disease |  |
| Read V3 (CTV3) | E00.. | Senile and presenile organic psychot conditions (& dementia) |  |
| Read V3 (CTV3) | E000. | Uncomplicated senile dementia |  |
| Read V3 (CTV3) | E001. | Presenile dementia |  |
| Read V3 (CTV3) | E0010 | Uncomplicated presenile dementia |  |
| Read V3 (CTV3) | E0011 | Presenile dementia with delirium |  |
| Read V3 (CTV3) | E0012 | Presenile dementia with paranoia |  |
| Read V3 (CTV3) | E0013 | Presenile dementia with depression |  |
| Read V3 (CTV3) | E001z | Presenile dementia NOS |  |
| Read V3 (CTV3) | E002. | Senile dementia with depressive or paranoid features |  |
| Read V3 (CTV3) | E0020 | Senile dementia with paranoia |  |
| Read V3 (CTV3) | E0021 | Senile dementia with depression |  |
| Read V3 (CTV3) | E002z | Senile dementia with depressive or paranoid features NOS |  |
| Read V3 (CTV3) | E003. | Senile dementia with delirium |  |
| Read V3 (CTV3) | E004. | Arteriosclerotic dementia (including [multi infarct dement]) |  |
| Read V3 (CTV3) | E0040 | Uncomplicated arteriosclerotic dementia |  |
| Read V3 (CTV3) | E0041 | Arteriosclerotic dementia with delirium |  |
| Read V3 (CTV3) | E0042 | Arteriosclerotic dementia with paranoia |  |
| Read V3 (CTV3) | E0043 | Arteriosclerotic dementia with depression |  |
| Read V3 (CTV3) | E004z | Arteriosclerotic dementia NOS |  |
| Read V3 (CTV3) | E00y. | (Oth senile/presen org psychoses) or (presbyophren psychos) |  |
| Read V3 (CTV3) | E00z. | Senile or presenile psychoses NOS |  |
| Read V3 (CTV3) | E011. | Korsakov psychosis |  |
| Read V3 (CTV3) | E0110 | Korsakov psychosis |  |
| Read V3 (CTV3) | E0111 | Korsakov's alcoholic psychosis with peripheral neuritis |  |
| Read V3 (CTV3) | E0112 | Wernicke-Korsakov syndrome |  |
| Read V3 (CTV3) | E011z | Alcohol amnestic syndrome NOS |  |
| Read V3 (CTV3) | E012. | Alcoholic dementia: [other] or [NOS] |  |
| Read V3 (CTV3) | E040. | Korsakoff's syndrome - non-alcoholic |  |
| Read V3 (CTV3) | E041. | Dementia in conditions EC |  |
| Read V3 (CTV3) | Eu00. | [X]Dementia in Alzheimer's disease | x |
| Read V3 (CTV3) | Eu001 | Dementia in Alzheimer's disease with late onset | x |
| Read V3 (CTV3) | Eu002 | [X]Dementia in Alzheimer's dis, atypical or mixed type | x |
| Read V3 (CTV3) | Eu00z | [X]Dementia in Alzheimer's disease, unspecified | x |
| Read V3 (CTV3) | Eu01. | Vascular dementia |  |
| Read V3 (CTV3) | Eu011 | [X]Dementia: [multi-infarct] or [predominantly cortical] |  |
| Read V3 (CTV3) | Eu01y | [X]Other vascular dementia |  |
| Read V3 (CTV3) | Eu01z | [X]Vascular dementia, unspecified |  |
| Read V3 (CTV3) | Eu02. | [X]Dementia in other diseases classified elsewhere |  |
| Read V3 (CTV3) | Eu020 | [X]Dementia in Pick's disease |  |
| Read V3 (CTV3) | Eu021 | [X]Dementia in Creutzfeldt-Jakob disease |  |
| Read V3 (CTV3) | Eu022 | [X]Dementia in Huntington's disease |  |
| Read V3 (CTV3) | Eu023 | [X]Dementia in Parkinson's disease |  |
| Read V3 (CTV3) | Eu02y | [X]Dementia in other specified diseases classif elsewhere |  |
| Read V3 (CTV3) | Eu02z | [X] Dementia: [unspecified] or [named variants (& NOS)] |  |
| Read V3 (CTV3) | Eu041 | [X]Delirium superimposed on dementia |  |
| Read V3 (CTV3) | F110. | Alzheimer's disease | x |
| Read V3 (CTV3) | F1100 | Dementia in Alzheimer's disease with early onset | x |
| Read V3 (CTV3) | F1101 | Dementia in Alzheimer's disease with late onset | x |
| Read V3 (CTV3) | F111. | Pick's disease |  |
| Read V3 (CTV3) | F11x7 | Cerebral degeneration due to Creutzfeldt-Jakob disease |  |
| Read V3 (CTV3) | F11x8 | Cerebral degeneration due to multifocal leucoencephalopathy |  |
| Read V3 (CTV3) | F21y2 | Binswanger's disease |  |
| Read V3 (CTV3) | Fyu30 | [X]Other Alzheimer's disease | x |
| Read V3 (CTV3) | Ub1T6 | Language disorder of dementia |  |
| Read V3 (CTV3) | X002m | Amyotrophic lateral sclerosis with dementia |  |
| Read V3 (CTV3) | X002w | Dementia |  |
| Read V3 (CTV3) | X002x | Dementia in Alzheimer's disease with early onset | x |
| Read V3 (CTV3) | X002y | Familial Alzheimer's disease of early onset | x |
| Read V3 (CTV3) | X002z | Non-familial Alzheimer's disease of early onset | x |
| Read V3 (CTV3) | X0030 | Dementia in Alzheimer's disease with late onset | x |
| Read V3 (CTV3) | X0031 | Familial Alzheimer's disease of late onset | x |
| Read V3 (CTV3) | X0032 | Non-familial Alzheimer's disease of late onset | x |
| Read V3 (CTV3) | X0033 | Focal Alzheimer's disease | x |
| Read V3 (CTV3) | X0034 | Frontotemporal dementia |  |
| Read V3 (CTV3) | X0035 | Pick's disease with Pick bodies |  |
| Read V3 (CTV3) | X0036 | Pick's disease with Pick cells and no Pick bodies |  |
| Read V3 (CTV3) | X0037 | Frontotemporal degeneration |  |
| Read V3 (CTV3) | X0039 | Frontal lobe degeneration with motor neurone disease |  |
| Read V3 (CTV3) | X003A | Lewy body disease |  |
| Read V3 (CTV3) | X003G | Progressive aphasia in Alzheimer's disease | x |
| Read V3 (CTV3) | X003H | Argyrophilic brain disease |  |
| Read V3 (CTV3) | X003I | Post-traumatic dementia |  |
| Read V3 (CTV3) | X003J | Punch drunk syndrome |  |
| Read V3 (CTV3) | X003K | Spongiform encephalopathy |  |
| Read V3 (CTV3) | X003L | Prion protein disease |  |
| Read V3 (CTV3) | X003M | Gerstmann-Straussler-Scheinker syndrome |  |
| Read V3 (CTV3) | X003P | Acquired immune deficiency syndrome dementia complex |  |
| Read V3 (CTV3) | X003R | Vascular dementia of acute onset |  |
| Read V3 (CTV3) | X003T | Subcortical vascular dementia |  |
| Read V3 (CTV3) | X003V | Mixed cortical and subcortical vascular dementia |  |
| Read V3 (CTV3) | X003W | Semantic dementia |  |
| Read V3 (CTV3) | X003X | Patchy dementia |  |
| Read V3 (CTV3) | X003Y | Epileptic dementia |  |
| Read V3 (CTV3) | X003l | Parkinson's disease - dementia complex on Guam |  |
| Read V3 (CTV3) | X00R0 | Presbyophrenic psychosis |  |
| Read V3 (CTV3) | X00R2 | Senile dementia |  |
| Read V3 (CTV3) | X00Rk | Alcoholic dementia NOS |  |
| Read V3 (CTV3) | X73mf | Creutzfeldt-Jakob disease agent |  |
| Read V3 (CTV3) | X73mj | Bovine spongiform encephalopathy agent |  |
| Read V3 (CTV3) | XE1Xr | Senile and presenile organic psychotic conditions |  |
| Read V3 (CTV3) | XE1Xs | Vascular dementia |  |
| Read V3 (CTV3) | XE1Xt | Other senile and presenile organic psychoses |  |
| Read V3 (CTV3) | XE1Xu | Other alcoholic dementia |  |
| Read V3 (CTV3) | XE1Z6 | [X]Unspecified dementia |  |
| Read V3 (CTV3) | XE1aG | Dementia (& [presenile] or [senile]) |  |
| Read V3 (CTV3) | Xa0lH | Multi-infarct dementia |  |
| Read V3 (CTV3) | Xa0sC | Frontal lobe degeneration |  |
| Read V3 (CTV3) | Xa0sE | Dementia of frontal lobe type |  |
| Read V3 (CTV3) | Xa1GB | Cerebral degeneration presenting primarily with dementia |  |
| Read V3 (CTV3) | Xa25J | Alcoholic dementia |  |
| Read V3 (CTV3) | Xa3ez | Other senile/presenile dementia |  |
| Read V3 (CTV3) | XaA1S | New variant of Creutzfeldt-Jakob disease |  |
| Read V3 (CTV3) | XaE74 | Senile dementia of the Lewy body type |  |
| Read V3 (CTV3) | XaIKB | Alzheimer's disease with early onset | x |
| Read V3 (CTV3) | XaIKC | Alzheimer's disease with late onset | x |
| Read V3 (CTV3) | XaKyY | [X]Lewy body dementia |  |
| Read V3 (CTV3) | XaLFf | Exception reporting: dementia quality indicators |  |
| Read V3 (CTV3) | XaLFo | Excepted from dementia quality indicators: Patient unsuitabl |  |
| Read V3 (CTV3) | XaLFp | Excepted from dementia quality indicators: Informed dissent |  |
| Read V3 (CTV3) | XaMAo | Prion protein markers for Creutzfeldt-Jakob disease |  |
| Read V3 (CTV3) | XaMFy | Dementia monitoring administration |  |
| Read V3 (CTV3) | XaMG0 | Dementia monitoring first letter |  |
| Read V3 (CTV3) | XaMGF | Dementia annual review |  |
| Read V3 (CTV3) | XaMGG | Dementia monitoring second letter |  |
| Read V3 (CTV3) | XaMGI | Dementia monitoring third letter |  |
| Read V3 (CTV3) | XaMGJ | Dementia monitoring verbal invite |  |
| Read V3 (CTV3) | XaMGK | Dementia monitoring telephone invite |  |
| Read V3 (CTV3) | XaMJC | Dementia monitoring |  |
| Read V3 (CTV3) | XaZWz | Participates in Butterfly Scheme for dementia |  |
| Read V3 (CTV3) | XaZX0 | Butterfly Scheme for dementia declined |  |
| Read V3 (CTV3) | XabVp | Sporadic Creutzfeldt-Jakob disease |  |
| Read V3 (CTV3) | XabtQ | Dementia medication review |  |
| Read V3 (CTV3) | Xaghb | Predominantly cortical dementia |  |
| Read V3 (CTV3) | Y000c | Dementia review done |  |
| Read V3 (CTV3) | Y1f1d | Dementia monitoring invitation |  |
| Read V3 (CTV3) | Y1f22 | Dementia monitoring invitation |  |
| Read V3 (CTV3) | Y1f98 | Quality and Outcomes Framework dementia quality indicator-related care invitation (procedure) |  |
| Read V3 (CTV3) | Y23fb | Mixed dementia |  |
| Read V3 (CTV3) | Y6230 | Creutzfeldt - Jakob disease |  |
| Read V3 (CTV3) | Y8180 | Other senile/presenile dement. |  |
| Read V3 (CTV3) | Y9086 | Senile dementia - simple type |  |
| Read V3 (CTV3) | Y9087 | Senile dementia-acute confused |  |
| Read V2 | Fyu3000 | [X]Other Alzheimer's disease | x |
| Read V2 | Eu01.11 | [X]Arteriosclerotic dementia |  |
| Read V2 | E004300 | Arteriosclerotic dementia with depression |  |
| Read V2 | Eu04100 | [X]Delirium superimposed on dementia |  |
| Read V2 | E002z00 | Senile dementia with depressive or paranoid fe... |  |
| Read V2 | Eu00013 | [X]Alzheimer's disease type 2 | x |
| Read V2 | Eu01y00 | [X]Other vascular dementia |  |
| Read V2 | Eu02y00 | [X]Dementia in other specified diseases classi... |  |
| Read V2 | Eu02z11 | [X] Presenile dementia NOS |  |
| Read V2 | Eu00200 | [X]Dementia in Alzheimer's dis atypical or mi... | x |
| Read V2 | E00..11 | Senile dementia |  |
| Read V2 | Eu02z13 | [X] Primary degenerative dementia NOS |  |
| Read V2 | E000.00 | Uncomplicated senile dementia |  |
| Read V2 | E002100 | Senile dementia with depression |  |
| Read V2 | E001000 | Uncomplicated presenile dementia |  |
| Read V2 | Eu01000 | [X]Vascular dementia of acute onset |  |
| Read V2 | Eu00012 | [X]Primary degen dementia Alzheimer's type p... | x |
| Read V2 | Eu00100 | [X]Dementia in Alzheimer's disease with late o... | x |
| Read V2 | E004.00 | Arteriosclerotic dementia |  |
| Read V2 | E004000 | Uncomplicated arteriosclerotic dementia |  |
| Read V2 | E001100 | Presenile dementia with delirium |  |
| Read V2 | E004z00 | Arteriosclerotic dementia NOS |  |
| Read V2 | Eu01300 | [X]Mixed cortical and subcortical vascular dem... |  |
| Read V2 | E004200 | Arteriosclerotic dementia with paranoia |  |
| Read V2 | E004.11 | Multi infarct dementia |  |
| Read V2 | F112.00 | Senile degeneration of brain |  |
| Read V2 | F110.00 | Alzheimer's disease | x |
| Read V2 | E00..12 | Senile/presenile dementia |  |
| Read V2 | E002.00 | Senile dementia with depressive or paranoid fe... |  |
| Read V2 | F110000 | Alzheimer's disease with early onset | x |
| Read V2 | E001300 | Presenile dementia with depression |  |
| Read V2 | E003.00 | Senile dementia with delirium |  |
| Read V2 | E001z00 | Presenile dementia NOS |  |
| Read V2 | E001200 | Presenile dementia with paranoia |  |
| Read V2 | Eu01200 | [X]Subcortical vascular dementia |  |
| Read V2 | Eu00011 | [X]Presenile dementiaAlzheimer's type | x |
| Read V2 | Eu02z14 | [X] Senile dementia NOS |  |
| Read V2 | E004100 | Arteriosclerotic dementia with delirium |  |
| Read V2 | Eu02z16 | [X] Senile dementia depressed or paranoid type |  |
| Read V2 | Eu00112 | [X]Senile dementiaAlzheimer's type | x |
| Read V2 | Eu00.00 | [X]Dementia in Alzheimer's disease | x |
| Read V2 | Eu00000 | [X]Dementia in Alzheimer's disease with early ... | x |
| Read V2 | F110100 | Alzheimer's disease with late onset | x |
| Read V2 | Eu02z00 | [X] Unspecified dementia |  |
| Read V2 | Eu01z00 | [X]Vascular dementia unspecified |  |
| Read V2 | E001.00 | Presenile dementia |  |
| Read V2 | Eu01100 | [X]Multi-infarct dementia |  |
| Read V2 | E002000 | Senile dementia with paranoia |  |
| Read V2 | Eu01111 | [X]Predominantly cortical dementia |  |
| Read V2 | Eu00113 | [X]Primary degen dementia of Alzheimer's type... | x |
| Read V2 | Eu00z11 | [X]Alzheimer's dementia unspec | x |
| Read V2 | 1461 | H/O: dementia |  |
| Read V2 | Eu00z00 | [X]Dementia in Alzheimer's disease unspecified | x |
| Read V2 | Eu01.00 | [X]Vascular dementia |  |
| Read V2 | Eu00111 | [X]Alzheimer's disease type 1 | x |

Read V3 codes were based on the following code list (ID 48c76cf8) from OpenCodelists:

<https://www.opencodelists.org/codelist/opensafely/dementia-complete/48c76cf8/#full-list>

Read V2 codes were based on code list PH859 / 1797 from the CALIBER phenotype library:

<https://phenotypes.healthdatagateway.org/phenotypes/PH859/version/1797/detail/>

**Supplemental Table 2: code list for AD ascertainment in FinnGen**

*Registry filters for endpoint ‘G6_AD_Strict’ (“Alzheimer’s disease, strict definition”) in FinnGen data release 13:*

| **Source (classification schema)** | **Code(s)** |
| --- | --- |
| Hospital discharge (ICD-10) | [F00.0*](https://icd.who.int/browse10/2016/en#/F00.0*), [F00.00*](https://icd.who.int/browse10/2016/en#/F00.00*), [F00.00*G30.0](https://icd.who.int/browse10/2016/en#/F00.00*G30.0), [F00.1*](https://icd.who.int/browse10/2016/en#/F00.1*), [F00.10*](https://icd.who.int/browse10/2016/en#/F00.10*), [F00.10*G30.1](https://icd.who.int/browse10/2016/en#/F00.10*G30.1), [G30.0](https://icd.who.int/browse10/2016/en#/G30.0), [G30.0+F00.00](https://icd.who.int/browse10/2016/en#/G30.0+F00.00), [G30.1](https://icd.who.int/browse10/2016/en#/G30.1), [G30.1+F00.10](https://icd.who.int/browse10/2016/en#/G30.1+F00.10) |
| Hospital discharge (ICD-9) | 3310 |
| Hospital discharge (ICD-8) | 29010 |
| Cause of death (ICD-10) | [F00.0*](https://icd.who.int/browse10/2016/en#/F00.0*), [F00.00*](https://icd.who.int/browse10/2016/en#/F00.00*), [F00.00*G30.0](https://icd.who.int/browse10/2016/en#/F00.00*G30.0), [F00.1*](https://icd.who.int/browse10/2016/en#/F00.1*), [F00.10*](https://icd.who.int/browse10/2016/en#/F00.10*), [F00.10*G30.1](https://icd.who.int/browse10/2016/en#/F00.10*G30.1), [G30.0](https://icd.who.int/browse10/2016/en#/G30.0), [G30.0+F00.00](https://icd.who.int/browse10/2016/en#/G30.0+F00.00), [G30.1](https://icd.who.int/browse10/2016/en#/G30.1), [G30.1+F00.10](https://icd.who.int/browse10/2016/en#/G30.1+F00.10) |
| Cause of death (ICD-9) | 3310 |
| Cause of death (ICD-8) | 29010 |
| KELA reimbursements (KELA) | 307 |
| Medicine purchases (ATC) | [F00.0*](https://icd.who.int/browse10/2016/en#/F00.0*), [F00.00*](https://icd.who.int/browse10/2016/en#/F00.00*), [F00.00*G30.0](https://icd.who.int/browse10/2016/en#/F00.00*G30.0), [F00.1*](https://icd.who.int/browse10/2016/en#/F00.1*), [F00.10*](https://icd.who.int/browse10/2016/en#/F00.10*), [F00.10*G30.1](https://icd.who.int/browse10/2016/en#/F00.10*G30.1), [G30.0](https://icd.who.int/browse10/2016/en#/G30.0), [G30.0+F00.00](https://icd.who.int/browse10/2016/en#/G30.0+F00.00), [G30.1](https://icd.who.int/browse10/2016/en#/G30.1), [G30.1+F00.10](https://icd.who.int/browse10/2016/en#/G30.1+F00.10) |

Cases required a minimum of three recorded events.

**Supplemental Table 3: code list for all-cause dementia ascertainment in FinnGen**

*Registry filters for endpoint ‘F5_Dementia’ (“Dementia”) in FinnGen data release 13:*

| **Source (classification schema)** | **Code(s)** |
| --- | --- |
| Hospital discharge (ICD-10) | [F00-F09](https://icd.who.int/browse10/2016/en#/F00-F09) |
| Hospital discharge (ICD-9) | 290 \| 3310 \| 4378A |
| Hospital discharge (ICD-8) | 290 |
| Cause of death (ICD-10) | [F00-F09](https://icd.who.int/browse10/2016/en#/F00-F09) |
| Cause of death (ICD-9) | 290 \| 3310 \| 4378A |
| Cause of death (ICD-8) | 290 |
| KELA reimbursements (KELA) | 307 |
| Medicine purchases (ATC) | N06D |

**Supplemental Figure 1: amyloid PET SUVr distribution in the A4 Study analytical sample (N=4,415)**

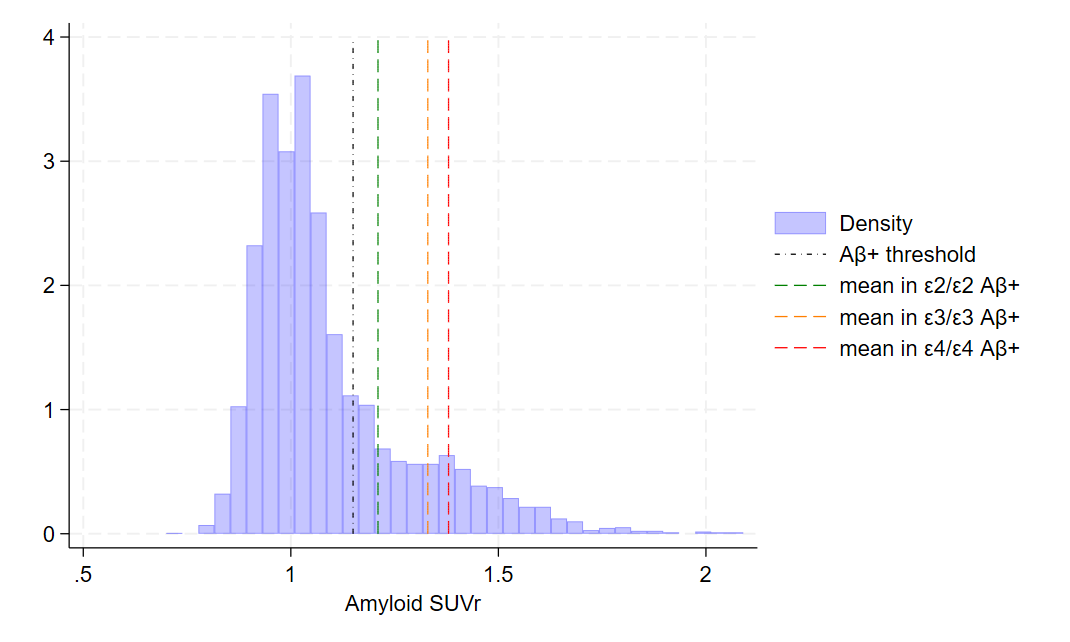
